# Divergent enlarged perivascular spaces volumes in early versus late age-of-onset Alzheimer’s disease

**DOI:** 10.1101/2023.08.01.23293514

**Authors:** Kyan Younes, Yann Cobbigo, Tori Tsuie, Earnest Wang, Amy Wolf, Renaud La Joie, David N Soleimani-Meigooni, Breton Asken, Duygu Tosun, Joel H Kramer, Adam R Ferguson, Bruce L Miller, Elizabeth C Mormino, Daniel Schwartz, Lisa C Silbert, Gil Rabinovici, Howard J Rosen, Fanny M Elahi

**Author notes:** **Corresponding Author:** Kyan Younes, MD., Address: 213 Quarry Rd, Palo Alto, California 94304.

## Abstract

**INTRODUCTION:** Enlarged perivascular spaces (EPVS) are considered a conduit for the brain’s waste clearance system. With aging, the brain’s ability to clear molecules is thought to decline, contributing to the retention of Alzheimer’s disease (AD) neuropathology. However, the role of EPVS in late-onset AD (LOAD) is complicated by co-morbidities. Early-onset AD (EOAD) offers a unique opportunity to understand the role of EPVS in AD.

**METHODS:** Automatically-segmented EPVS volumes in biomarker-confirmed EOAD (*n*=58), LOAD (*n*=43), and age-matched controls (*n*=60) were correlated with amyloid and tau PET and cognition. Linear regression models were used.

**RESULTS:** In LOAD, higher EPVS volumes were associated with better memory and functional performance. However, this association was not observed in EOAD. Additionally, higher tau was linked to increased EPVS in LOAD, but not in EOAD.

**DISCUSSION:** EOAD and LOAD demonstrate distinct associations between EPVS, AD hallmarks, and cognition, suggesting differences in EPVS’s role in these AD subtypes, necessitating further investigation.

## BACKGROUND

Perivascular spaces (PVS; a.k.a., Virchow-Robin Spaces) are cerebrospinal fluid-filled spaces that surround the cerebral blood vessels as they extend from the subarachnoid space into the brain parenchyma.^1^ In the past, enlarged PVS (EPVS) detected through neuroimaging were typically considered normal anatomical changes or unrelated to pathological brain aging and degeneration as they were commonly observed in cognitively unimpaired individuals.^2^ However, over the past decade, studies have linked EPVS to various conditions such as cerebrovascular disease and vascular contributions to dementia,^3^ neuroinflammation,^4^ and demyelination.^5^ The relationship between EPVS and Alzheimer’s disease (AD) remains somewhat unclear. Some studies have shown higher centrum semiovale EPVS in AD,^2^ but others showed no difference in EPVS between AD and healthy controls.^6–10^ Additionally, certain studies have suggested that basal ganglia EPVS are more prominent in vascular dementia,^3^ while centrum semiovale EPVS may be associated with cerebral amyloid angiopathy.^11^

EPVS is increasingly recognized as a potential marker of impaired glymphatic system function, responsible for clearing the brain’s metabolites and circulating immune cells, which may contribute to the build-up of AD neuropathology.^12^ Effective clearance of the brain’s biological byproducts is crucial to its health.^1^ The glymphatic system is a multi-cellular network connected with perivascular spaces that promote cerebrospinal fluid motion from the subarachnoid spaces, into the perivascular spaces, then into the surrounding brain parenchyma, followed by routing into the perivenular spaces, on its way into the jugular veins for brain clearance. This lifelong fluid flux clears metabolites and cells from the brain, retaining brain homeostasis.^1,45–47^However, the interpretation of EPVS in individuals with AD is challenging due to several factors. These factors include variations in EPVS volumes among individuals with and without the disease, differences in dementia stage, manual EPVS quantification methods (e.g., manual counts on an ordinal scale), or AD inclusion criteria (e.g., clinical consensus versus biomarker-supported).

Sporadic early onset AD (EOAD) is characterized by the presence of Aβ plaques and tau neurofibrillary tangles, similar to late-onset AD (LOAD), which is defined to occur after the age of 65 years. EOAD is diagnosed when the disease develops before the age of 65 years, without any autosomal dominant AD mutations, and tends to present more commonly with atypical non-amnestic forms of AD, such as executive, language, or visuospatial presentations, compared to the more amnestic presentation often seen in LOAD. Despite some shared commonalities, EOAD is believed to have a purer and more intense manifestation of AD neuropathology and is influenced by different polygenic factors compared to LOAD.^13–21^ In contrast, LOAD is more likely to be affected by aging-related processes such as limbic age-related TDP-43 encephalopathy, hippocampal sclerosis, and cerebrovascular disease.^13^ These distinctions in neuropathological features and underlying genetic influences between EOAD and LOAD contribute to the complexity in understanding the role of EPVS in AD.

In this study, we aimed to address the existing ambiguity and mixed findings regarding the role of EPVS in AD by comparing EPVS in EOAD and LOAD. Understanding the role of EPVS in these subtypes could provide valuable insights into the pathophysiology of AD, particularly considering the higher tau burden observed in EOAD compared to LOAD. To achieve this, we investigated the relationships between EPVS and various markers of AD progression, including amyloid and tau positron emission tomography (PET) signals, volumetric MRI cortical atrophy, neuropsychological evaluations, and functional measures. EPVS measures were derived using a semi-automatic method based on MRI data in participants with EOAD, LOAD, and demographically matched controls. Additionally, all participants underwent comprehensive clinical phenotyping and assessment of AD biomarkers. By examining EPVS in these AD subtypes and their correlations with other AD-related markers, we hope to gain a deeper understanding of EPVS’s potential role in the pathogenesis of AD. This research has the potential to contribute significantly to our knowledge of the disease and may have implications for future therapeutic approaches targeting EPVS and its impact on AD progression.

## METHODS

### Participants selection

Participants in this study were recruited and assessed at the University of California, San Francisco, Memory and Aging Center and Alzheimer’s Disease Research Center. The diagnosis was established through a comprehensive assessment and a consensus of behavioral neurologist, neuropsychologist, and nursing staff.^22^ We included individuals who met clinical criteria for mild cognitive impairment^23^ or dementia due to AD,^24^ had brain MRI, and positive amyloid PET with visits between 2016 and 2020 (n = 101). We used a cutoff of 65 years old for patient’s first diagnosis to determine if a patient belonged to the EOAD (*n* = 58) or the LOAD (*n* = 43) groups. A group of age-matched amyloid PET negative healthy aging controls from the MAC Hillblom Healthy Aging Network (*n* = 60, 23 participants younger than 65 and 37 older than 65 years old) was also included. Patients or caregivers provided informed consent following procedures aligned with the Declaration of Helsinki, and the study was approved by the UCSF Committee for Human Research.

### Functional, cognitive, and behavioral assessments

Patients underwent a comprehensive multidisciplinary assessment that included functional and neuropsychological measures (**Table 1**). A functional assessment was done through a semi-structured interview with the patient’s co-participant using the Clinical Dementia Rating Scale (CDR). Composite measures were used to summarize neuropsychological performance on several paper and pencil measures. Raw test scores were transformed to z-scores and then averaged across the subtests included in the composite.^25^ Memory composite included verbal and visual episodic memory tested with the California Verbal Learning Test and a 10-minute delayed free recall of the Benson complex figure recall, respectively. Visuospatial composite included the Benson complex figure copy and the Number Location subtest of the Visual Object Space Perception battery. Executive composite included backward digit span, modified trail making, Stroop color-naming test (inhibition trial), lexical fluency (D-words in one minute), and design fluency. Language composite included 15-item Boston Naming Test and animal fluency.

**Table 1.** Demographics, APOE, cognition, and function in the healthy controls (younger and older controls) and Alzheimer’s disease (Early Onset Alzheimer’s Disease and Late Onset Alzheimer’s Disease)

|  | Younger<br>healthy control | Older healthy<br>control | All healthy<br>controls | Early onset<br>Alzheimer's | Late onset<br>Alzheimer's | All Alzheimer's |
| --- | --- | --- | --- | --- | --- | --- |
|  | Age < 65 | Age >65 |  | Age < 65 | Age >65 |  |
| Count | 23 | 37 | 60 | 48 | 43 | 91 |
| Age, mean (sd) | 58 (4.1)b,e | 81 (4.9)b, f, c, d | 72 (12)a | 58 (4.8) b, d | 75 (6.1) b, f, d | 66.3 (10.1)a |
| Handedness, (right, non-right) | 20, 3 | 34, 3 | 54, 6 | 41, 7 | 38, 5 | 79, 12 |
| Sex, (male, female) | 10, 13 | 19, 34 | 29, 47 | 18, 30b | 25, 18b | 43, 48 |
| Education in years, mean (SD) | 16.4 (4) | 17.6 (1.9) d | 17.2 (2.9)a | 15.2 (4.3) d | 16.8 (3.6) | 15.9 (4.1)a |
| Ethnicity |  |  |  |  |  |  |
| White n, % | 15 (65) | 28 (75) | 43 (71) | 40 (83) | 41 (95) | 81 (89) |
| Asian n, % | 2 (8) | 1 (2) | 3 (5) | 1 (2) | 0 (0) | 1 (1) |
| Hispanic n, % | 0 (0) | 1 (2) | 1 (1) | 1 (2) | 0 (0) | 1 (1) |
| Black n, % | 3 (13) | 1 (3) | 4 (7) | 2 (4) | 0 (0) | 2 (2) |
| American Indian or Alaska Native n, % | 2 (8) | 2 (5) | 4 (7) | 0 (0) | 0 (0) | 0 (0) |
| Other n, % | 0 (0) | 2 (5) | 2 (3) | 3 (6) | 0 (0) | 3 (3) |
| Unknown n, % | 0 (0) | 0 (0) | 0 (0) | 0 (0) | 4 (9) | 4 (4) |
| APOE |  |  |  |  |  |  |
| Two E4 alleles | 2 (8) | 0 (0) | 2 (3) | 4 (8) | 6 (14) | 10 (11) |
| One E4 allele | 6 (26) | 5 (14) | 11 (18) | 19 (40) | 18 (42) | 37 (41) |
| No E4 alleles | 14 (61) | 30 (81) | 44 (73) | 25 (52) | 20 (47) | 45 (49) |
| Function and Cognition |  |  |  |  |  |  |
| CDR score, mean (sd), max = 3 | 0.04 (0.14) g, e | 0.014 (0.08) d, f | 0.03 (0.11)a | 0.77 (0.51) d, g | 0.86 (0.78) e, f | 1.32 (3.74)a |
| CDR Box Score, mean (sd), max = 18 | 0.11 (0.37) g, e | 0.04 (0.14) d, f | 0.07 (0.25)a | 4.42 (2.83) d, g | 3.88 (2.44) e, f | 4.11 (4.12)a |
| **MMSE, mean (sd), max = 30** | 29.63 (0.67) g, e | 28.39 (1.55) d, f | 28.68 (1.49)a | 21.37 (6.43) d, g | 24.13 (3.84) e, f | 23.31 (5.91)a |
| **Memory composite, mean (sd)** | 0.05 (0.90) g, e | 0.54 (0.82) d, f | 0.36 (0.88)a | -2.62 (1.61) d, g | -2.18 (1.28) e, f | -2.41 (1.46)a |
| **Executive composite, mean (sd)** | 0.28 (0.45) g, e | 0.22 (0.50) d, f | 0.24 (0.48)a | -2.12 (1.42)b,d,g | -1.41 (1.42)b,e,f | -1.77(1.38)a |
| **Visuospatial composite, mean (sd)** | 0.11 (0.45) g, e | 0.24 (0.63) d, f | 0.20 (0.56)a | -3.56 (4.01)b,d,g | -1.00 (2.81)b,e,f | -2.32 (3.68)a |
| **Language composite, mean (sd)** | 0.33 (1.10) g, e | 0.12 (0.45) d, f | 0.23 (0.82)a | -2.28 (2.36) d, g | -2.20 (2.19) e, f | -2.24 (2.26)a |

a: CO is different from AD
b: EOAD is different from LOAD
c: Younger CO is different from Older CO
d: EOAD different from Older CO
e: LOAD different from Younger CO
f: LOAD different from Older control
g: EOAD different from Younger CO

### Image Acquisition

#### MRI image acquisition

Participants underwent a whole-brain imaging on a 3 Tesla Siemens Trio Total imaging matrix, with a 12-channel head coil, or Prisma Fit, with a 64-channel head coil, magnetic resonance imaging (MRI) system. T1-weighted images were acquired with the Magnetization Prepared Rapid Gradient Echo (MP-RAGE) sequences (Sagittal slice orientation; 240 × 256 matrix, 160 slices, voxel size = 1.0 × 1.0 × 1.0 mm^3^; TR = 2300 ms; TE 2.98 ms; flip angle = 9°).

#### PET image acquisition

PET scans were performed at the Lawrence Berkeley National Laboratory on a Siemens Biograph six Truepoint PET/CT scanner in 3D acquisition mode. A low-dose CT/transmission scan was performed for attenuation correction prior to all scans. Pittsburgh Compound-B (PIB) and Flortaucipir (FTP) PET were synthesized and acquired as previously described.^26^ Amyloid-PET data was analyzed at 50–70 minutes after injection of approximately 15 mCi of PIB, and tau-PET data was analyzed at 80–100 minutes after injection of approximately 10 mCi of FTP. PET data were reconstructed in 5-min frames using an ordered subset expectation maximization algorithm with weighted attenuation. Images were smoothed with a 4 mm Gaussian kernel with scatter correction and evaluated prior to analysis for patient motion and adequacy of statistical counts.

### Image processing

#### MRI image processing

Before any prepossessing of the images, all T1-weighted images were visually inspected for quality control. T1-weighted images underwent bias field correction using N3 algorithm, the segmentation was performed using SPM12 (Wellcome Trust Center for Neuroimaging http://www.fil.ion.ucl.ac.uk/spm, London, UK) unified segmentation.^27^ A group template was generated from the segmented tissue probability maps by non-linear registration template generation using Large Deformation Diffeomorphic Metric Mapping framework.^28^ Global white matter hyperintensity (WMH) volume was obtained using native space FLAIR and T1-weighted images as previously described.^29^

#### EPVS image processing

An unsupervised segmentation algorithm was used to quantify EPVS burden within WM tissue. Total EPVS burden was quantified within a compact 90% binarized white matter mask, created in the T1-weighted native space using SPM. Corresponding T2-weighted images was rigidly registered into T1-weighted patient space. The EPVS were segmented following a semi-automated process,^29^ the curvilinear structures were isolated using Sato’s filter.^30^ A density-based clustering algorithm iterated on the segmented mask independently labeled unique EPVS, providing an estimate of total and local count and volume.^31^ Images were visually inspected for quality and signal in the ventricle, lacune, or sulcal regions were manually removed.

#### PET image processing

PET frames were realigned, averaged, and co-registered to the MPRAGE images using Statistical Parametric Mapping (SPM) Version 12. Standardized uptake value ratios (SUVR) were calculated for the 50–70-min post-injection interval of PIB using mean activity in the cerebellar cortex gray matter as the reference region, and for the 80–100-min post-injection interval of FTP using mean activity in the inferior cerebellar cortex gray matter as the reference region.^32^ To estimate global amyloid burden, a global PIB index SUVR was calculated for each patient using a composite of frontal, parietal, temporal and cingulate regions known to show high PIB binding in AD.^33^ SUVR values were calculated then converted to Centiloid (CL) units using a previously validated conversion equation.^34,35^ Amyloid-PET positivity was based on quantitative PIB SUVR cutoff of 1.21,^36,37^ which has been validated against post-mortem amyloid burden. All EOAD and LOAD patients were amyloid-PET positive, and all healthy controls were amyloid-PET negative per inclusion criteria. To estimate tau burden, mean values of FTP PET SUVR were extracted from the whole cortex, the precuneus, and bilateral temporal meta-ROI (weighted average of fusiform gyrus, parahippocampal gyrus, entorhinal cortex, inferior temporal cortex, middle temporal cortex, and amygdala).

### Genotyping

To assess APOE status, genomic DNA was extracted from peripheral blood using standard protocols. APOE genotyping was carried out by real-time PCR on a LightCycler® 480 System using Taqman SNP Genotyping Assays. Genotyping results were available on 148/161 subjects (48/58 EOAD,43/43 LOAD, 57 healthy controls).

### Statistical analyses

Tests of normality for all continuous data in AD and Controls were conducted with the Shapiro-Wilk test. Means for continuous variables were compared with the Student’s t or the Mann-Whitney U tests, where appropriate. Baseline characteristics were compared using the χ2 or Fisher exact test for categorical variables.

To understand the mutual variance affecting EPVS in the entire sample including Controls and AD, we conducted a series of univariate correlations between EPVS volume and each of age, white matter volume, gray matter volume, intracranial volume, brain parenchymal fraction (used as surrogate marker for atrophy and calculated as: (gray matter + white matter) / total intracranial volume), and white matter hyperintensity volume. White matter volume was the only variable that correlated with EPVS volume, albeit weakly (*r* = 0.23, *p value* = 0.004), and therefore all subsequent analyses were corrected for white matter volume **(Supplementary Figure 1)**. ANOVA was used to compare EPVS volume in all four groups (EOAD, LOAD, Younger Controls, and Older Controls) and was Bonferroni adjusted for multiple comparisons.

To investigate whether the relationship of EPVS volume corrected for white matter volume with each of age, gray matter volume adjusted for intracranial volume, brain parenchymal fraction, and white matter hyperintensities differed between AD and Controls (**Supplementary Figure 2**), we performed a series of multiple linear regression models including the interaction of diagnostic group by EPVS volume (i.e., group (AD or Control) * EPVS volume).

To investigate the differences in EPVS volume in EOAD and LOAD by each of tau or amyloid, we conducted linear regression models including the interaction term of diagnostic group by each of these variables (i.e., EPVS volume ∼ age + sex + education + group + tau+ group * tau; EPVS volume ∼ age + sex + education + group + amyloid + group * amyloid) (**Supplementary Table 1 and Figure 2**).

To determine the association between EPVS volume and the cognitive and functional outcomes, a series of full factorial design multivariable linear regression models including diagnostic group by EPVS volume interaction terms were examined. Adjusting for age, sex, and education, we compared the relationship between EPVS corrected for white matter volume in Controls and AD (EOAD and LOAD combined) and each of clinical dementia rating scale sum of boxes (i.e., CDR-SB ∼ age + sex + education + group + EPVS volume + group * EPVS volume), memory (i.e., memory composite ∼ age + sex + education + group + EPVS volume + group * EPVS volume), executive function (i.e., executive composite ∼ age + sex + education + group + EPVS volume + group * EPVS volume), visuospatial i.e., visuospatial composite ∼ age + sex + education + group + EPVS volume + group * EPVS volume), and language (i.e., language composite ∼ age + sex + education + group + EPVS volume + group * EPVS volume). Next, we investigated the interaction between AD group (EOAD vs LOAD) and EPVS volume on cognitive and functional performance (**Figure 3, Supplementary Table 2, Supplementary Figure 3**).

Finally, to explore whether the effect of EPVS on tau was conditional on the CDR sum of boxes, we ran an exploratory full factorial design three-way interaction of (i.e., tau ∼ age + education + sex + EPVS_WM_log + CDR-Sum of Boxes + Group (EOAD_LOAD) + EPVS_WM_log * CDR-Sum of Boxes * Group (EOAD_LOAD)) **Supplementary Table 3**. To visualize a three-way interaction, we plotted the marginal effects of the interaction terms (**Figure 4)**.

All analyses were performed using R Version 4.0.2 with RStudio versions 1.3.1056. Results were considered to be statistically significant at *p value <* .05. Statistical assumptions for all analyses were tested and met. All observations were independent by study design. For ANOVA and linear regression analyses, response variables data were continuous. Visual inspection of Q-Q plots revealed that error terms were approximately normally distributed.

## RESULTS

### Demographic and clinical differences

Demographic and clinical data are presented in **Table 1**. As a whole, the sample was predominantly white (81%) and highly educated (mean education = 16.7 years). There were no statistically significant demographic differences found between EOAD and LOAD or AD as a whole with Controls in handedness or race/ethnicity. There were no differences between AD and Controls in sex but within the AD group, there was more women in the EOAD group. The healthy control group had more years of completed education than the AD group and the LOAD group had more years of education than the EOAD group. There were no significant differences in prevalence of *APOE4* genotype between EOAD and LOAD. All subsequent statistical analyses were corrected for sex and education. Since age cut-offs were distinct for EOAD and LOAD, as part of grouping, models with and without adjusting for age were performed.

### No differences in EPVS volume between groups

EPVS volume and white matter hyperintensity volume were right skewed in all cohorts (**Figure 1a**) and therefore they were log transformed in the subsequent analyses to normalize their distributions. There were no statistically significant differences in the raw uncorrected or the white matter volume corrected EPVS volume between AD and Controls, or between EOAD, LOAD, younger Controls, and older Controls (**Figure 1b**). There was also no statistically significant interaction between diagnostic group and EPVS volume on age, gray matter volume, white matter hyperintensity, and brain parenchymal fraction (**Supplementary Results, Supplementary Figure 2**).

**Figure 1.**
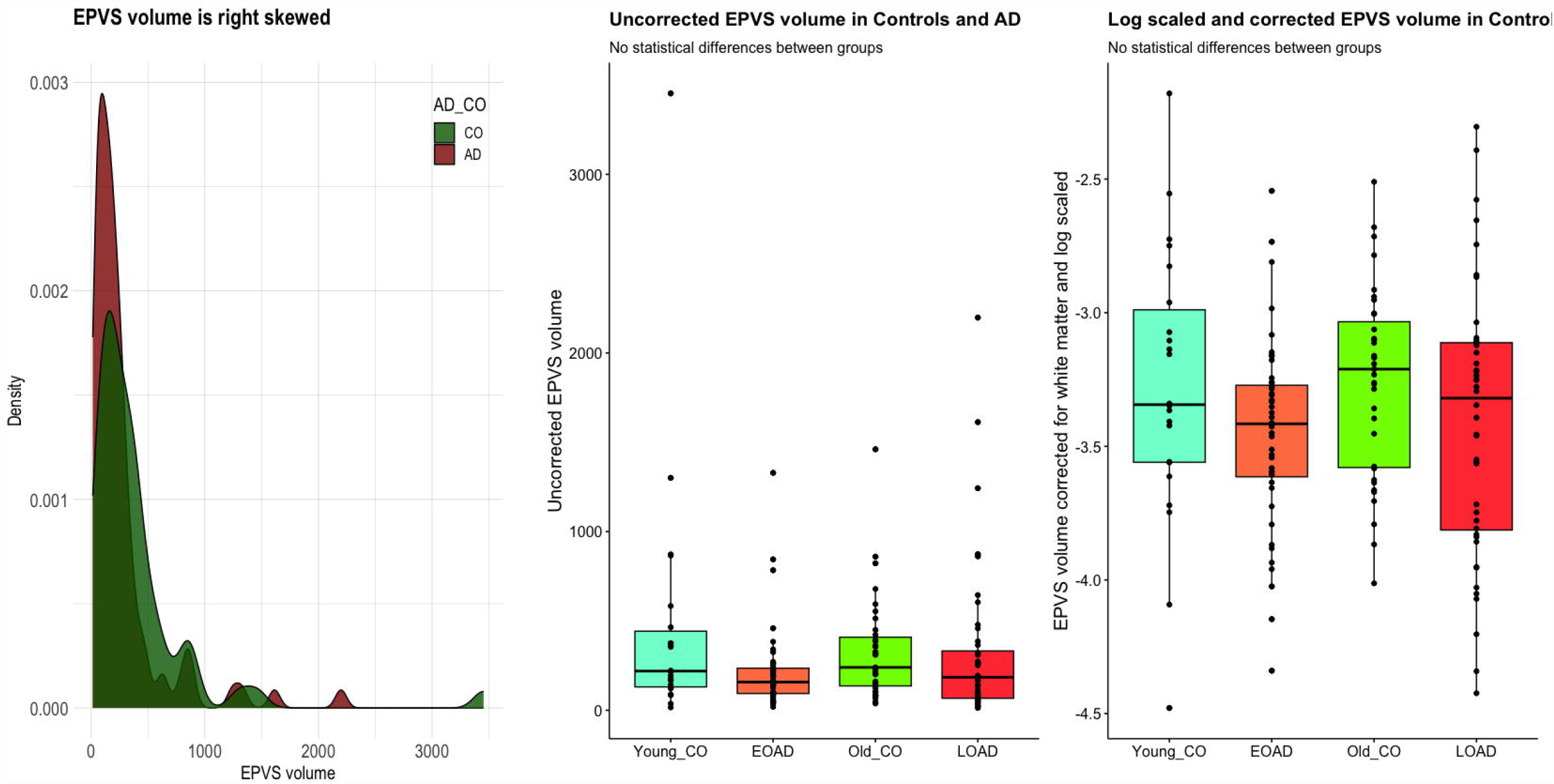
Left: EPVS volume density distribution; EPVS volume is right skewed and therefore it was log scaled to normalize the distribution. Middle: Boxplot showing uncorrected EPVS volume across EOAD; LOAD, younger healthy aging CO, older healthy aging CO. There were no statistically significant differences between EOAD, LOAD, Younger CO, or Older CO. Abbreviations: CO: Controls; EOAD: Early Onset Alzheimer’s Disease. EPVS: Enlarged Perivascular Space; LOAD: Late Onset Alzheimer’s Disease; ROI: Region of Interest.

### EPVS relationship to amyloid and tau

In LOAD, higher global cortical tau SUVR, temporal meta ROI SUVR, precuneus SUVR, and amyloid centeloids were associated with higher EPVS volume. This relationship was not seen in EOAD (**Figure 2 and Supplementary Table 1**). There was a significant interaction of group (EOAD or LOAD) * Temporal meta-FTP (*b* = .50, *p value* = .02), group (EOAD vs LOAD) * Global FTP SUVR (b = .52, *p value* = 0.04), and marginally significant group (EOAD vs LOAD) * PIB centiloids (b = .43, *p value* = .06) on EPVS volume corrected for white matter volume. AD group (EOAD vs LOAD) * precuneus FTP was not statistically significance (b = .47, *p value* = .42). These results show that higher tau and amyloid burdens were associated with higher EPVS volume in LOAD but not in EOAD, suggesting a divergent role for EPVS in LOAD and EOAD pathophysiology or a role for differing co-pathologies contributing to the association of AD neuropathology with EPVS.

**Figure 2.**
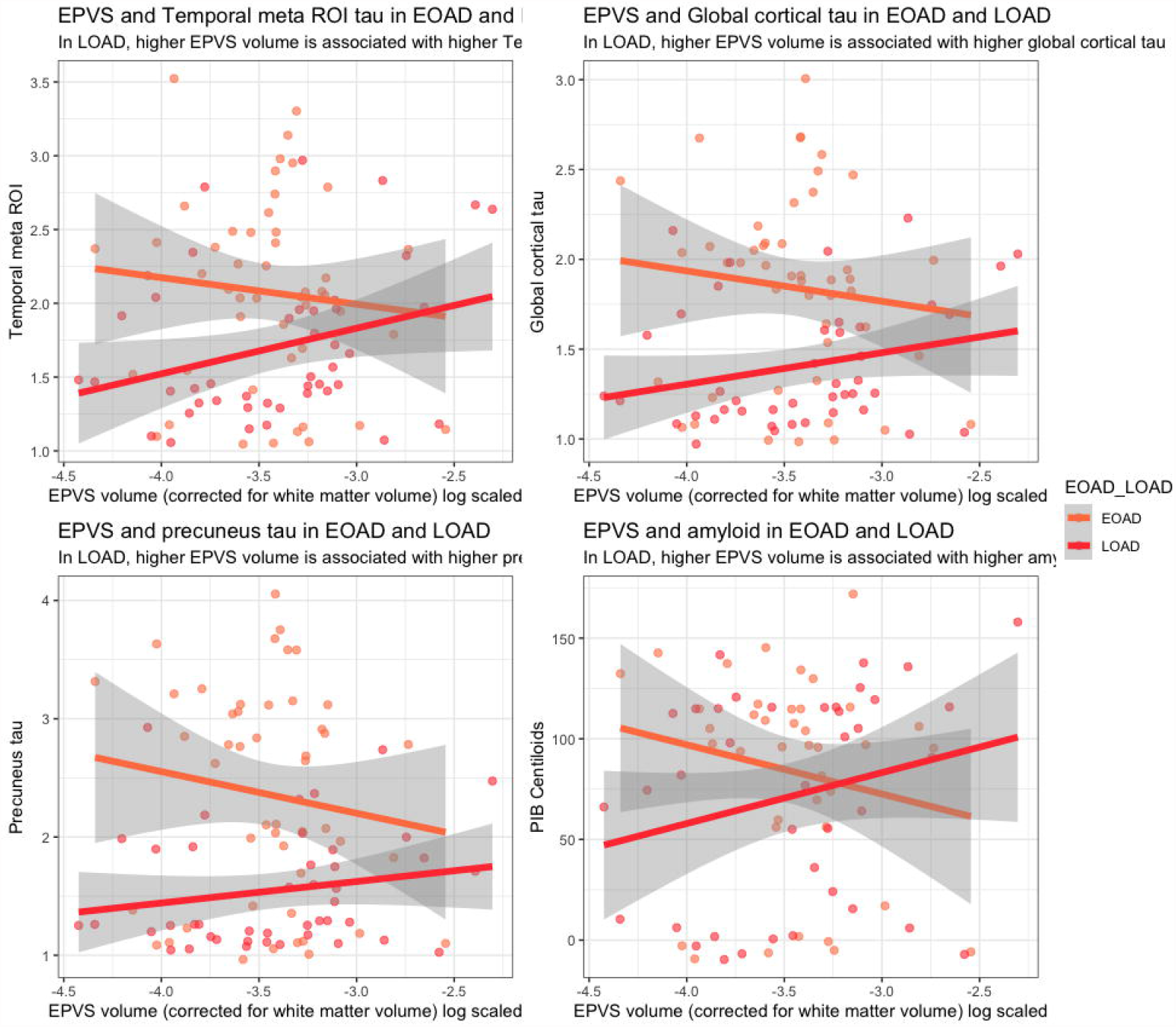
EPVS correlates with tau and amyloid in LOAD. Figure shows the relationship between EPVS volume corrected for white matter volume and log scaled with each of global cortical tau, meta temporal tau, precuneus tau and amyloid on EPVS volume. In LOAD, higher global cortical tau, meta temporal tau, and amyloid were associated with higher EPVS volume. See Supplementary Table 1 for statistical details. Abbreviations: EOAD: Early Onset Alzheimer’s Disease. EPVS: Enlarged Perivascular Space; LOAD: Late Onset Alzheimer’s Disease; ROI: Region of Interest.

### EPVS relationship to cognition and function

We first compared the entire AD cohort (EOAD and LOAD combined) to Controls. Higher EPVS volume was associated with better memory in AD (*b* = 0.19, *p* = 0.02) and the interaction term EPVS * Group (AD vs Controls) was statistically significant (*b* = -0.26, *p* = 0.05). There were no association or significant interaction between EPVS and Group (AD vs Controls) on executive, visuospatial, or language composites. On the functional scale (CDR-SoB), there were no significant differences in EPVS volumes between AD and Controls, however there was a trend for higher EPVS volume correlating with better functional state on CDR-SoB (*b* = -0.13, *p* = 0.09) (**Figure 3, Supplementary Table 2**).

**Figure 3.**
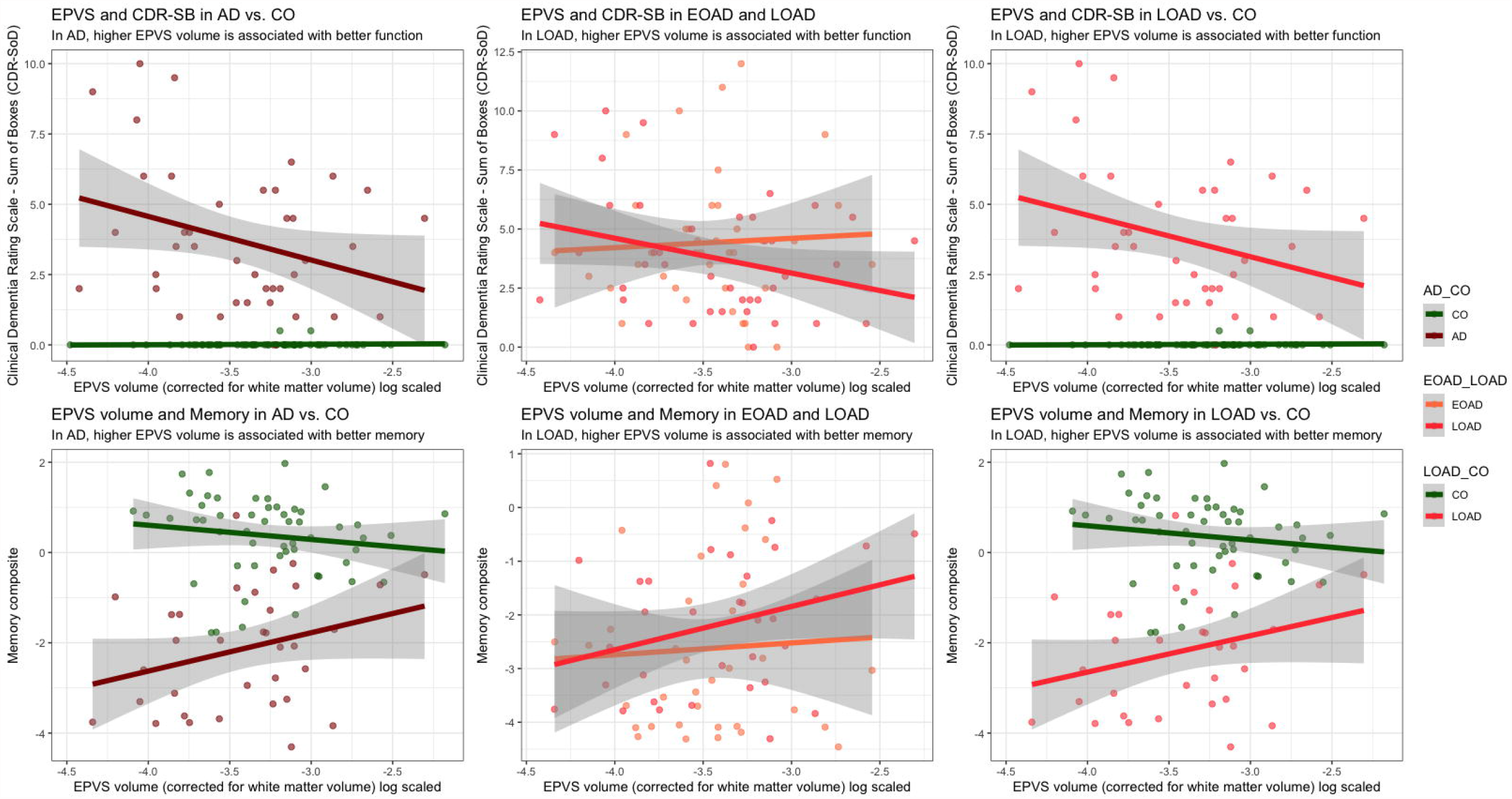
EPVS association with functional and cognitive scores in EOAD, LOAD and CO. Left Panel shows that in AD higher EPVS is associated with better functional (CDR-SB), and memory scores. The Middle Panel shows that within AD, the differences seen on the Left Panel is driven by the LOAD group. The Right Panel shows that in LOAD higher EPVS is associated with better functional (CDR-SB), and memory scores compared to CO. There were no differences on visuospatial, executive, and language composite scores (Supplementary Figure 3). See Supplementary Table Supplementary Table 2 for statistical details. Abbreviations: CDR-SB: Clinical Dementia Rating Scale – Sum of Boxes; EOAD: Early Onset Alzheimer’s Disease. EPVS: Enlarged Perivascular Space; LOAD: Late Onset Alzheimer’s Disease; ROI: Region of Interest.

Next, we wanted to test whether EOAD and LOAD had differences, similar to the differences noted for AD neuropathologies. We investigated the interaction between AD group (EOAD vs LOAD) and EPVS volume on cognitive and functional performance, and found no significant interactions on any cognitive outcomes. There was a trend for higher EPVS volume correlating with lower CDR sum of boxes in LOAD (*b*= -0.41, *p*= 0.08) (**Supplementary Figure 3, Supplementary Table 2)**.

Because of the abovementioned findings (i.e., significant AD/Control and marginally significant LOAD/EOAD interactions with EPVS volume on Memory and CDR sum of boxes), we compared LOAD to Controls on those two measures (**Figure 3 and Supplementary Table 2**). There were significant main effects and interactions for EPVS * Group (LOAD vs Controls) on memory composite (*b* = 0.31, *p value*

= 0.01) and CDR sum of boxes (*b* = -1.49, *p value* = 0.03)—indicating an association of EPVS in LOAD but not EOAD in the direction of higher EPVS volume associating with better memory and functional performances.

### Three-Way-Interaction EPVS, Box Score, and Tau PET

To investigate the association between EPVS and tau across clinical stages of disease—marked by CDR sum-of-boxes scores—in each of EOAD and LOAD, we ran a three-way interaction which showed distinct patterns. The three-way interaction term was statistically significant (**Figure 4, Supplementary Table 3**). In LOAD, when the disease is mild (low CDR-SB), higher EPVS volume is associated with relatively unchanged tau levels (i.e., elevated EPVS volume keeps tau levels stable). As the disease severity increases, elevated EPVS volume is associated with higher tau level in a CDR-SB in dose-dependent fashion (moderate disease, elevated EPVS associated with moderate slope of tau increase and severe disease elevated EPVS associated with higher slope of tau accumulation). In other words, as the disease advances tau accumulation for the same amount of EPVS increases. This relationship is different in EOAD, in mild disease higher EPVS volume is associated with higher tau accumulation whereas in severe disease higher EPVS is associated with lower tau. Given the exploratory nature of this analysis, we ran another three-way interaction to investigate the association between tau and CDR-SB for the degree of EPVS volume (**Supplementary Figure 3)**. This interaction was not statistically significant, however it showed that LOAD patients with the highest EPVS volume, tended to have the lowest tau and CDR sum of boxes early in the disease course consistent with the previous analyses and suggesting that in LOAD subjects with higher EPVS volume perform better functionally than subjects with low EPVS volume for the same level of tau as measured by tau PET.

**Figure 4.**
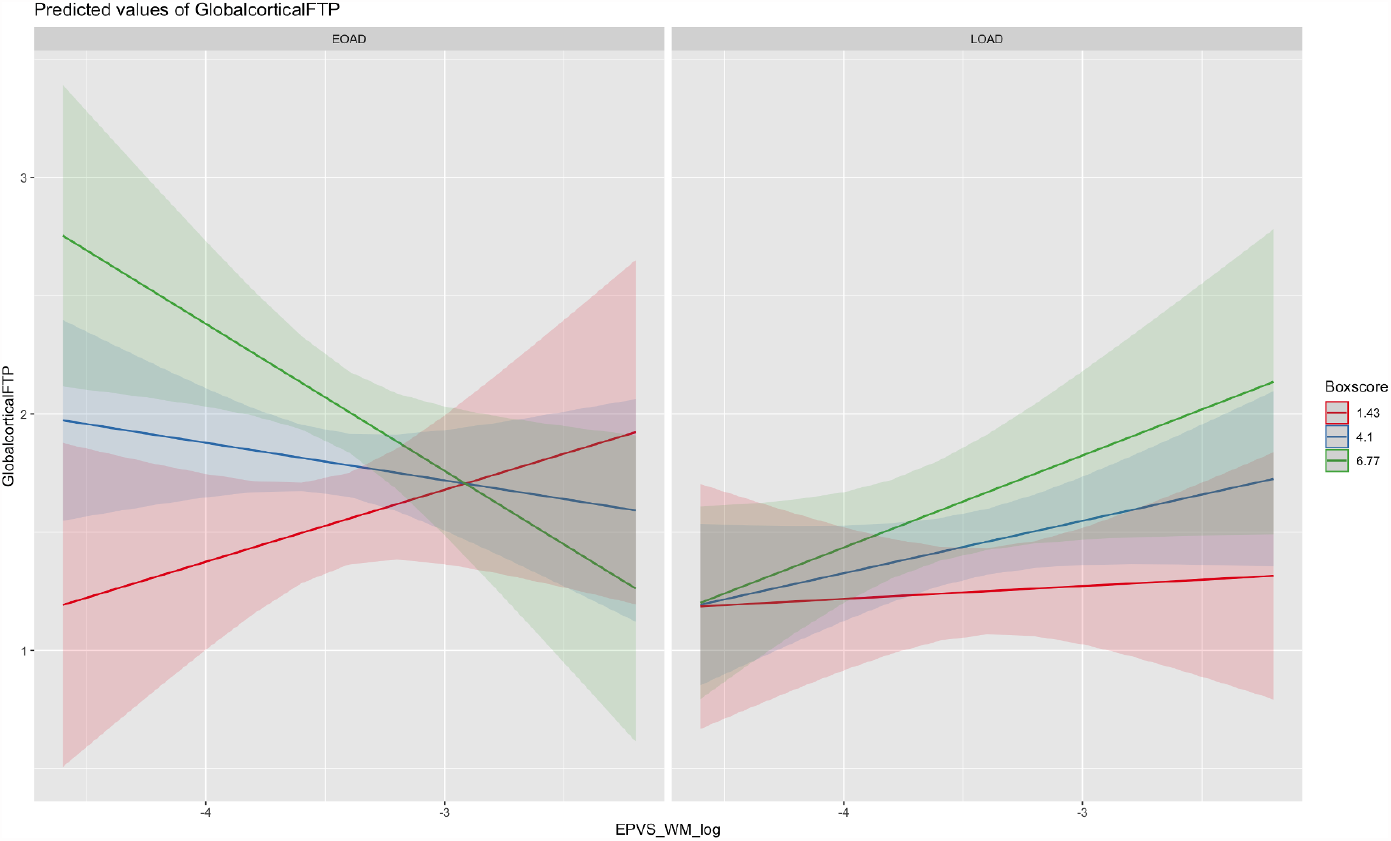
EPVS volume relationship with tau differs based on disease stage and age of onset. For lowest CDR tertile (red), in EOAD higher EPVS is associated with higher tau, in later disease (green) higher EPVS is associated lower tau. Whereas in LOAD, in early disease higher EPVS is not associated with increased tau, in later disease stages higher EPVS is associated with higher tau and the slope is “dose-dependent” on disease severity (i.e., as the disease advances the rate of tau accumulation for the same amount of EPVS increases). See Supplementary Table 3 for statistical details. Abbreviations: CDR-SB: Clinical Dementia Rating Scale – Sum of Boxes; EOAD: Early Onset Alzheimer’s Disease. EPVS: Enlarged Perivascular Space; LOAD: Late Onset Alzheimer’s Disease; ROI: Region of Interest.

## DISCUSSION

This study investigates the associations of MRI-visible EPVS with brain function using cognitive and functional assessments, as well as AD-relevant biomarkers, such as cortical grey-matter volumes, brain burdens of amyloid, and tau in early and late age of onset AD. Our findings reveal distinct EPVS associations in EOAD versus LOAD. Specifically, EPVS correlated with higher tau burden but better brain function in LOAD. This suggests that higher EPVS volumes could serve as a putative compensatory mechanism in the presence of pathology. Although MRI-visible EPVS could represent various cellular and extracellular components, they are commonly believed to play a role in brain clearance. It is conceivable that dysfunction in clearance mechanisms is more commonly related to LOAD pathophysiology, which occurs at a later age and has a higher burden of vascular pathology. On the other hand, EOAD may be less influenced by vascular dysfunction. Animal and cellular models have supported a role for EPVS in amyloid clearance in AD.^38–40^ Pre-dating the description of the glymphatic system, EPVS imaging studies suggested a higher number of EPVS in MCI and dementia.^41–44^ However, methodological choices (such as including visual counting and ordinal scales, not accounting for total white matter volume, and including heterogenous cohorts without AD-biomarker status) limited the interpretation of EPVS in AD pathology. Our findings provide a potential explanation for the ambiguous relationship of EPVS and AD in the literature by showing a dynamic role for EPVS in AD pathophysiology, which varies based on the clinical and pathological disease stages, including early and late age-of-onset subtypes. Further research is needed to fully elucidate the significance of EPVS in AD and its potential as a therapeutic target.

We formulated two explanations for the observed associations between EPVS and accumulated tau and amyloid proteins in LOAD. The first hypothesis is referred to as the clearance dysfunction hypothesis. This hypothesis states that there may be upstream pathologies hindering clearance, resulting in impaired flow and leading to enlarged EPVS volumes and higher amyloid and tau burdens. The second hypothesis, the compensatory role hypothesis, is that EPVS may expand to increase the clearance rate of amyloid and tau in response to increased levels of these proteins. Since our data is cross-sectional, we cannot determine causality or differentiate between these explanations. However, the fact that the individuals with higher EPVS volume exhibit better cognitive and functional performance despite elevated amyloid and tau levels suggests that the compensatory role hypothesis merits further investigation. We speculate that this finding is a reflection of variable upstream phenomena, whereby vascular comorbidities and clearance mechanisms play a more important role in the pathophysiology of LOAD, while EOAD is less reliant on these risk factors.

Our data show that on quantitative evaluation of EPVS volume, there were no differences between healthy aging Controls and individuals affected with AD. In AD patients and Controls, there were no differences in the associations between EPVS volume and age, gray matter volume, white matter hyperintensity volume, or brain parenchymal fraction. There were however differences in associations of EPVS volumes and cognitive and functional scores based on whether subjects had AD or not. Consistently, Sepehrband et al automatically measured EPVS in a biomarker-proven AD cohort found an inverse correlation between EPVS and tau PET in the superior mesial temporal lobe.^48^ In a post-mortem MRI study, EPVS burden was associated with pre-mortem faster decline in visuospatial abilities, lower levels of semantic memory and visuospatial abilities at the time of death in patients with dementia and without dementia.^49^ It is possible that the existing literature reflects the different stages in a dynamic role that EPVS play in brain health and disease. As a potential neuroimaging marker in dementia-causing diseases, such as AD, more studies are needed to understand the shifting role that EPVS may play in brain function. EPVS may reflect an active (enlarged PVS) vs inactive (not enlarged PVS) clearance system. Alternatively, specific locations of EPVS may reflect obstruction due to accumulated molecules and cells.

Mixed neuropathology is present in almost all older brains, in both patients with no cognitive difficulties and patients with neurodegenerative disease pathologies and clinical deficits.^50^ This suggests that a slower brain clearance may occur in aging and neurodegeneration. Postmortem examination of EPVS has been associated with both AD neuropathology and vascular pathologies, including astrogliosis.^51^ A study that measured EPVS volume in young adults suggested that EPVS volume is subject to changes depending on body mass index and genetic profile.^52^ In our cohort, LOAD patients with elevated tau but high EPVS volume performed better on functional and memory scales, possibly reflecting differences in the pathophysiology of EAOD versus LOAD and suggesting a dynamic role for EPVS in the disease stage.

EPVS volume and CSF dynamics may be factors that interact with amyloid and tau in the AD pathophysiological cascade and this interaction is variable by disease stage. AD is a dynamic disease where multiple factors are interacting over the course of many years. Therefore, multi-pronged interventions that consider the stage of each biological process and the interaction between these processes might be necessary for successful therapeutics. For instance, therapies that could enhance the brain clearance system might be employed early in the disease stage when EPVS are still large, and function and cognition are relatively preserved. Future longitudinal studies should investigate the stability of EPVS volume overtime, their longitudinal relationship with amyloid and tau as well as other proteionopathies such as alpha-synuclein and TDP-43.

The limitations of this study include the difficulty establishing causality with regards to the role and implications of EPVS in the context of AD pathology, despite the multimodal imaging approach. Longitudinal data are crucial to better understand the dynamic nature of EPVS^52^ and its relationship with AD progression. Secondly, although we provide data correlating EPVS volume with amyloid and tau burden, we base the role of EPVS in the waste clearance function on preceding animal models literature. Neuroimaging methods that directly measure brain waste clearance in humans are needed. Unfortunately, this study also lacks ethnic, racial, and demographic diversity and larger more inclusive datasets are needed to understand ethnic and racial variability in EPVS volume. Additionally, our sample size is small for the three-way interaction and larger sample sizes are needed. Larger, longitudinal, and more inclusive studies that include multimodal imaging and investigate the underlying pathophysiology of EPVS changes will help expand on our findings.

Therapeutic approaches targeting misfolded protein clearance in AD may face challenges due to a potential vicious cycle: worsening glymphatic clearance leading to increased protein accumulation, which in turn further impairs glymphatic clearance. Noninvasive methods to study the glymphatic system in the human brain will be instrumental in understanding this dynamic process and its implications for potential therapies. Our current multimodal neuroimaging study is the first to include EOAD patients and examines the relationship between EPVS, AD pathological hallmarks, and cognition. Although our study cannot establish causality between EPVS volumes and various outcomes, we present compelling evidence of different associations in early versus late age of onset AD. This highlights the importance of considering the dynamic role of EPVS in the pathophysiology of AD and encourages further investigations to fully comprehend its potential as a neuroimaging marker in dementia-causing diseases.

## Data Availability

The data for this study are available on request

## Abbreviations

AD: Alzheimer’s Disease
BPF: Brain Parenchymal Fraction
EPVS: Enlarged perivascular spaces
EOAD: Early Onset Alzheimer’s Disease
FTP: flortaucipir
LOAD: Late Onset Alzheimer’s Disease
PiB: Pittsburgh Compound-B.

## ACKNOWLEDGMENTS

The authors thank the research patients and their families for the time and effort they dedicated to research at UCSF’s Memory and Aging Center. We thank Dr Michael Greicius (Department of Neurology and Neurological Sciences, Stanford University, Stanford, CA, USA) for helpful discussions.

## CONSENT STATEMENT

All human subjects or caregivers provided informed consent following procedures aligned with the Declaration of Helsinki.

## REFERENCES

1. Iliff JJ, Wang M, Liao Y, et al. A Paravascular Pathway Facilitates CSF Flow Through the Brain Parenchyma and the Clearance of Interstitial Solutes, Including Amyloid. Sci Transl Med. 2012;4(147):147ra111–147ra111. doi:10.1126/scitranslmed.3003748

2. Smeijer D, Ikram MK, Hilal S, Kamran Ikram M, Hilal S. Enlarged Perivascular Spaces and Dementia: A Systematic Review. J Alzheimer’s Dis. 2019;72(1):247–256. doi:10.3233/JAD-190527

3. Doubal FN, MacLullich AMJ, Ferguson KJ, Dennis MS, Wardlaw JM. Enlarged Perivascular Spaces on MRI Are a Feature of Cerebral Small Vessel Disease. Stroke. 2010;41(3):450–454. doi:10.1161/STROKEAHA.109.564914

4. Rouhl RPW, Van Oostenbrugge RJ, Knottnerus ILH, Staals JEA, Lodder J. Virchow-Robin spaces relate to cerebral small vessel disease severity. J Neurol. 2008;255(5):692–696. doi:10.1007/s00415-008-0777-y

5. Patankar TF, Mitra D, Varma A, Snowden J, Neary D, Jackson A. Dilatation of the Virchow-Robin space is a sensitive indicator of cerebral microvascular disease: Study in elderly patients with dementia. Am J Neuroradiol. 2005;26(6):1512–1520.

6. Patankar T, Widjaja E, Chant H, et al. Relationship of Deep White Matter Hyperintensities and Cerebral Blood Flow in Severe Carotid Artery Stenosis.

7. Shams S, Martola J, Charidimou A, et al. Topography and Determinants of Magnetic Resonance Imaging (MRI)-Visible Perivascular Spaces in a Large Memory Clinic Cohort. J Am Heart Assoc. 2017;6(9). doi:10.1161/JAHA.117.006279

8. Chen W, Song X, Zhang Y. Assessment of the virchow-robin spaces in Alzheimer disease, mild cognitive impairment, and normal aging, using high-field MR imaging. Am J Neuroradiol. 2011;32(8):1490–1495. doi:10.3174/ajnr.A2541

9. Ramirez J, Berezuk C, McNeely AA, Gao F, McLaurin JA, Black SE. Imaging the Perivascular Space as a Potential Biomarker of Neurovascular and Neurodegenerative Diseases. Cell Mol Neurobiol. 2016;36(2):289–299. doi:10.1007/s10571-016-0343-6

10. Banerjee G, Kim HJ, Fox Z, et al. MRI-visible perivascular space location is associated with Alzheimer’s disease independently of amyloid burden. Brain. 2017;140(4):1107–1116. doi:10.1093/brain/awx003

11. Charidimou A, Hong YT, Jäger HR, et al. White Matter Perivascular Spaces on Magnetic Resonance Imaging: Marker of Cerebrovascular Amyloid Burden? Stroke. 2015;46(6):1707–1709. doi:10.1161/STROKEAHA.115.009090

12. Goldman SA, Benveniste H, Deane R, et al. A Paravascular Pathway Facilitates CSF Flow Through the Brain Parenchyma and the Clearance of Interstitial Solutes, Including Amyloid. Sci Transl Med. 2012;4(147):147ra111–147ra111. doi:10.1126/scitranslmed.3003748

13. Spina S, la Joie R, Petersen C, et al. Comorbid neuropathological diagnoses in early vs late-onset Alzheimer’s disease. medRxiv. 2020;(2021). doi:10.1101/2020.10.14.20213017

14. Harper L, Bouwman F, Burton EJ, et al. Patterns of atrophy in pathologically confirmed dementias: A voxelwise analysis. J Neurol Neurosurg Psychiatry. 2017;88(11):908–916. doi:10.1136/jnnp-2016-314978

15. Loy CT, Schofield PR, Turner AM, Kwok JBJ. Genetics of dementia. Lancet. 383(9919):828–840. doi:http://dx.doi.org/10.1016/S0140-6736(13)60630-3

16. Ducharme S, Dickerson BC. The Neuropsychiatric Examination of the Young-Onset Dementias. Psychiatr Clin North Am. 2015;38(2):249–264. doi:10.1016/j.psc.2015.01.002

17. Papageorgiou SG, Kontaxis T, Bonakis A, Kalfakis N, Vassilopoulos D. Frequency and causes of early-onset dementia in a tertiary referral center in Athens. Alzheimer Dis Assoc Disord. 2009;23(4):347–351. http://www.ncbi.nlm.nih.gov/entrez/query.fcgi?cmd=Retrieve&db=PubMed&dopt=Citation&list_uids=19568157

18. Reid W, Broe G, Creasey H, et al. Age at onset and pattern of neuropsychological impairment in mild early-stage Alzheimer disease. A study of a community-based population. Arch Neurol. 1996;53(10):1056–1061. http://www.ncbi.nlm.nih.gov/entrez/query.fcgi?cmd=Retrieve&db=PubMed&dopt=Citation&list_uids=8859068

19. Janssen JC, Beck JA, Campbell TA, et al. Early onset familial Alzheimer’s disease: Mutation frequency in 31 families. Neurology. 2003;60(2):235–239. http://www.ncbi.nlm.nih.gov/entrez/query.fcgi?cmd=Retrieve&db=PubMed&dopt=Citation&list_uids=12552037

20. Ossenkoppele R, Cohn-Sheehy BI, La Joie R, et al. Atrophy patterns in early clinical stages across distinct phenotypes of Alzheimer’s disease. Hum Brain Mapp. 2015;36(11):4421–4437. doi:10.1002/hbm.22927

21. Carmona S, Hardy J, Guerreiro R. Chapter 26 - The genetic landscape of Alzheimer disease. In: Geschwind DH, Paulson HL, Klein C, eds. Handb Clin Neurol. Vol 148. Elsevier; 2018:395–408. doi:https://doi.org/10.1016/B978-0-444-64076-5.00026-0

22. Rosen HJ, Hartikainen KM, Jagust W, et al. Utility of clinical criteria in differentiating frontotemporal lobar degeneration (FTLD) from AD. Neurology. 2002;58(11):1608–1615. http://www.ncbi.nlm.nih.gov/entrez/query.fcgi?cmd=Retrieve&db=PubMed&dopt=Citation&list_uids=12058087

23. Albert MS, Dekosky ST, Dickson D, et al. The diagnosis of mild cognitive impairment due to Alzheimer’s disease: Recommendations from the National Institute on Aging and Alzheimer’s Association workgroup. Alzheimers Dement. Published online 2011. http://www.ncbi.nlm.nih.gov/entrez/query.fcgi?cmd=Retrieve&db=PubMed&dopt=Citation&list_uids=21514249

24. McKhann GM, Knopman DS, Chertkow H, et al. The diagnosis of dementia due to Alzheimer’s disease: Recommendations from the National Institute on Aging and the Alzheimer’s Association workgroup. Alzheimers Dement. Published online 2011. http://www.ncbi.nlm.nih.gov/entrez/query.fcgi?cmd=Retrieve&db=PubMed&dopt=Citation&list_uids=21514250

25. Staffaroni AM, Brown JA, Casaletto KB, et al. The Longitudinal Trajectory of Default Mode Network Connectivity in Healthy Older Adults Varies As a Function of Age and Is Associated with Changes in Episodic Memory and Processing Speed. J Neurosci. 2018;38(11):2809–2817. doi:10.1523/JNEUROSCI.3067-17.2018

26. La Joie R, Visani A V., Lesman-Segev OH, et al. Association of APOE4 and Clinical Variability in Alzheimer Disease With the Pattern of Tau- and Amyloid-PET. Neurology. 2021;96(5):e650–e661. doi:10.1212/WNL.0000000000011270

27. Ashburner J, Friston KJ. Unified segmentation. Neuroimage. 2005;26(3):839–851. doi:10.1016/j.neuroimage.2005.02.018

28. Ashburner J, Friston KJ. Diffeomorphic registration using geodesic shooting and Gauss-Newton optimisation. Neuroimage. 2011;55(3):954–967. doi:10.1016/j.neuroimage.2010.12.049

29. Ithapu V, Singh V, Lindner C, et al. Extracting and summarizing white matter hyperintensities using supervised segmentation methods in Alzheimer’s disease risk and aging studies. Hum Brain Mapp. 2014;35(8):4219–4235. doi:10.1002/hbm.22472

30. Sato Y, Nakajima S, Shiraga N, et al. Three-dimensional multi-scale line filter for segmentation and visualization of curvilinear structures in medical images. Med Image Anal. 1998;2(2):143–168. doi:https://doi.org/10.1016/S1361-8415(98)80009-1

31. Boespflug EL, Schwartz DL, Lahna D, et al. MR imaging-based multimodal autoidentification of perivascular spaces (mMAPS): Automated morphologic segmentation of enlarged perivascular spaces at clinical field strength. Radiology. 2018;286(2):632–642. doi:10.1148/radiol.2017170205

32. Maass A, Landau S, Horng A, et al. Comparison of multiple tau-PET measures as biomarkers in aging and Alzheimer’s disease. Neuroimage. 2017;157(December 2016):448–463. doi:10.1016/j.neuroimage.2017.05.058

33. Rabinovici GD, Furst AJ, Alkalay A, et al. Increased metabolic vulnerability in early-onset Alzheimer’s disease is not related to amyloid burden. Brain. 2010;133(Pt 2):512–528. doi:10.1093/brain/awp326

34. Klunk WE, Koeppe RA, Price JC, et al. The Centiloid project: Standardizing quantitative amyloid plaque estimation by PET. Alzheimer’s Dement. 2015;11(1):1-15.e4. doi:10.1016/j.jalz.2014.07.003

35. Lesman-Segev OH, La Joie R, Stephens ML, et al. Tau PET and multimodal brain imaging in patients at risk for chronic traumatic encephalopathy. NeuroImage Clin. 2019;24(October):102025. doi:10.1016/j.nicl.2019.102025

36. Rabinovici GD, Rosen HJ, Alkalay A, et al. Amyloid vs FDG-PET in the differential diagnosis of AD and FTLD. Neurology. 2011;77(23):2034–2042. doi:10.1212/WNL.0b013e31823b9c5e

37. Villeneuve S, Rabinovici GD, Cohn-Sheehy BI, et al. Existing Pittsburgh Compound-B positron emission tomography thresholds are too high: statistical and pathological evaluation. Brain. 2015;138(Pt 7):2020–2033. doi:10.1093/brain/awv112

38. Zuroff L, Daley D, Black KL, Koronyo-Hamaoui M. Clearance of cerebral Abeta in Alzheimer’s disease: reassessing the role of microglia and monocytes. Cell Mol Life Sci. 2017;74(12):2167–2201. doi:10.1007/s00018-017-2463-7

39. Tarasoff-Conway JM, Carare RO, Osorio RS, et al. Clearance systems in the brain - Implications for Alzheimer disease. Nat Rev Neurol. 2015;11(8):457–470. doi:10.1038/nrneurol.2015.119

40. Natale G, Limanaqi F, Busceti CL, et al. Glymphatic System as a Gateway to Connect Neurodegeneration From Periphery to CNS. Front Neurosci. 2021;15(February):1–13. doi:10.3389/fnins.2021.639140

41. Smeijer D, Ikram MK, Hilal S. Enlarged Perivascular Spaces and Dementia: A Systematic Review. J Alzheimer’s Dis. 2019;72(1):247–256. doi:10.3233/JAD-190527

42. J MacLullich AM, Wardlaw JM, Ferguson KJ, et al. Enlarged perivascular spaces are associated with cognitive function in healthy elderly men. J Neurol Neurosurg Psychiatry. 2004;75(11):1519–1523. doi:10.1136/jnnp.2003.030858

43. Hilal S, Tan CS, Adams HHH, et al. Enlarged perivascular spaces and cognition. Neurology. 2018;91(9):e832–e842. doi:10.1212/WNL.0000000000006079

44. Bown CW, Carare RO, Schrag MS, Jefferson AL. Physiology and Clinical Relevance of Enlarged Perivascular Spaces in the Aging Brain. Published online 2021. doi:10.1212/WNL.0000000000013077

45. Iliff JJ, Nedergaard M. Is there a cerebral lymphatic system? Stroke. 2013;44(SUPPL. 1):93–95. doi:10.1161/STROKEAHA.112.678698

46. Jessen NA, Sofie A, Munk F, et al. HHS Public Access. 2016;40(12):198–202. doi:10.1016/j.jad.2015.06.025.Associations

47. Fang Y, Gu LY, Tian J, et al. MRI-visible perivascular spaces are associated with cerebrospinal fluid biomarkers in Parkinson’s disease. Aging (Albany NY). 2020;12(24):25805–25818. doi:10.18632/aging.104200

48. Sepehrband F, Barisano G, Sheikh-Bahaei N, et al. Alteration of perivascular spaces in early cognitive decline. bioRxiv Neurosci. 2020;(323). doi:10.1101/2020.01.30.927350

49. Javierre-Petit C, Schneider JA, Kapasi A, et al. Neuropathologic and Cognitive Correlates of Enlarged Perivascular Spaces in a Community-Based Cohort of Older Adults. Stroke. 2020;(September):1–9. doi:10.1161/strokeaha.120.029388

50. Boyle PA, Yu L, Wilson RS, Leurgans SE, Schneider JA, Bennett DA. Person-specific contribution of neuropathologies to cognitive loss in old age. Ann Neurol. 2018;83(1):74–83. doi:10.1002/ana.25123

51. Boespflug EL, Simon MJ, Leonard E, et al. Targeted Assessment of Enlargement of the Perivascular Space in Alzheimer’s Disease and Vascular Dementia Subtypes Implicates Astroglial Involvement Specific to Alzheimer’s Disease. J Alzheimer’s Dis. 2018;66(4):1587–1597. doi:10.3233/JAD-180367

52. Barisano G. Body mass index, time of day, and genetics affect perivascular spaces in the white matter Authors□: Affiliations□: Corresponding Author□: 2020;1.

