## Supplementary Materials for "Divergent enlarged perivascular spaces volumes in early versus late age-of-onset Alzheimer’s disease"

### Differential association of EPVS with clinical outcomes in early-onset versus late-onset Alzheimer`s Disease

#### Supplementary results:

##### No differences in EPVS volume between groups:

EPVS volume and white matter hyperintensity volume were right skewed in all cohorts and therefore they were log transformed in the subsequent analyses to normalize the distribution. There were no statistically significant differences in the raw uncorrected or the white matter volume corrected EPVS volume between AD and Controls, or between EOAD, LOAD, Younger Controls, and Older Controls (**Figure 1**). There was also no statistically significant interaction between diagnostic group and EPVS volume on each of age, gray matter volume, white matter hyperintensity, brain parenchymal fraction.

There was also no statistically significant effect of age on EPVS in AD or Controls, or in EOAD, LOAD, Younger Controls, or Older Controls with or without correction for white matter volume. The interaction terms Group (AD vs Controls)*Age ($p$ = 0.06), Group (AD vs Controls)*gray matter volume ($p$ = 0.76), Group (AD vs Controls)*white matter volume ($p$ = 0.66), Group (AD vs Controls)*white matter hyperintensity volume ($p$ = 0.26), and Group (AD vs Controls)*brain parenchymal fraction ($b$ = 0.03, $p$ = 0.08) were not significantly assoiated with EPVS volume (**Figure 1 and Supplementary Figure 2**).

Within the AD group, the interaction terms AD group (EOAD vs LOAD)*gray matter volume (b = -0.01, p = *0.53*), AD group (EOAD vs LOAD)*white matter hyperintensity volume (b= 0.01, $p$ = 0.99), AD group (EOAD vs LOAD)*Brain Parenchymal Fraction (b= 0.03, $p$ = 0.88) did not significantly associate with EPVS volume (**Supplementary Figure 2**).

**Supplementary Table 1.** Linear regression models output examining the relationship between AD pathological hallmarks (amyloid or tau) and EPVS in AD. Abbreviations: CDR-SB: Clinical Dementia Rating Scale – Sum of Boxes; EOAD: Early Onset Alzheimer`s Disease. EPVS: Enlarged Perivascular Space; LOAD: Late Onset Alzheimer`s Disease; ROI: Region of Interest.

**Supplementary Table 1a: Uncorrected for demographics.**

EPVS_WM_log ~ PET marker + EOAD_LOAD + PET marker * EOAD_LOAD

| Model | EPVS_WM_log | EPVS_WM_log | EPVS_WM_log | EPVS_WM_log |
|---|---|---|---|---|
| (Intercept) | -3.31***<br>(-0.23) | -3.34***<br>(-0.21) | -3.33***<br>(-0.17) | -3.35***<br>(-0.14) |
| Globalcortical FTP | -0.08<br>(-0.12) | | | |
| Group: LOAD | -0.62<br>(-0.36) | -0.58<br>(-0.31) | -0.37<br>(-0.28) | -0.26<br>(-0.18) |
| GlobalcorticalFTP*Group: LOAD | 0.45*<br>(-0.22) | | | |
| Temporal meta ROI FTP | | -0.05<br>(-0.1) | | |
| Temporal meta ROI FTP*Group: LOAD | | 0.35*<br>(-0.16) | | |
| Precuneus FTP | | | -0.05<br>(-0.07) | |
| PrecuneusFTP*Group: LOAD | | | 0.24<br>(-0.15) | |
| PIB Centiloids | | | | - 0.003<br>(0.001) |
| PIBCentiloids*Group: LOAD | | | | 0.003<br>(0.001) |

**Supplementary Table 1b: Corrected for age and education.**

EPVS_WM_log ~ age + education + PET marker + EOAD_LOAD + PET marker * Group (EOAD_LOAD)

| | EPVS_WM_log | EPVS_WM_log | EPVS_WM_log | EPVS_WM_log |
|---|---|---|---|---|
| Intercept | -3.94***<br>(-0.68) | -3.88***<br>(-0.6) | -3.86***<br>(-0.63) | -3.41***<br>(-0.67) |
| Age | -0.01<br>(-0.01) | -0.01<br>(-0.01) | -0.01<br>(-0.01) | 0.00<br>(-0.01) |
| Education | 0.00<br>(-0.01) | 0.00<br>(-0.01) | 0.00<br>(-0.01) | -0.01<br>(-0.01) |
| Globalcortical FTP | -0.05<br>(-0.12) | | | |
| Group: LOAD | -0.73<br>(-0.38) | -0.71*<br>(-0.34) | -0.49<br>(-0.31) | -0.32<br>(-0.23) |
| GlobalcorticalFTP*Group: LOAD | 0.43<br>(-0.23) | | | |
| Temporal meta ROI FTP | | -0.05<br>(-0.1) | | |
| Temporal meta ROI FTP*Group: LOAD | | 0.35*<br>(-0.16) | | |
| Precuneus FTP | | | -0.04<br>(-0.07) | |
| PrecuneusFTP*Group: LOAD | | | 0.24<br>(-0.15) | |
| PIB Centiloids | | | | - 0.003<br>(0.001) |
| PIBCentiloids*Group: LOAD | | | | 0.003<br>(0.001) |

**Supplementary Table 2** Linear regression models output examining the relationship between cognitive and functional outcomes and EPVS in AD. Abbreviations: CDR-SB: Clinical Dementia Rating Scale – Sum of Boxes; EOAD: Early Onset Alzheimer`s Disease. EPVS: Enlarged Perivascular Space; LOAD: Late Onset Alzheimer`s Disease; ROI: Region of Interest.

**Supplementary Table 2a** Cognition/function ~ education + EPVS_WM_log + Group (CO vs. LOAD) + EPVS_WM_log*Group (CO vs. LOAD)

| Model - LOAD:CO | CDR-SoB *B* (Std. Error) | Memory *B* (Std. Error) | Executive *B* (Std. Error) | Visuospatial *B* (Std. Error) | Language *B* (Std. Error) |
|---|---|---|---|---|---|
| Intercept | 0.05 (-1.86) | -0.37 (-1.45) | -0.85 (-1.15) | 2.67 (-2.52) | 0.25 (-2.11) |
| Education | 0 (-0.06) | -0.02 (-0.05) | 0.02 (-0.04) | -0.14 (-0.09) | -0.08 (-0.08) |
| EPVS_WM_log | 0.02 (-0.49) | -0.33 (-0.36) | -0.22 (-0.28) | 0.02 (-0.63) | -0.43 (-0.53) |
| Group: LOAD | -1.37 (-2.37) | 1.42 (-1.86) | -1.99 (-1.36) | -0.62 (-3.21) | 0.21 (-2.66) |
| EPVS_WM_log*Group: LOAD | **-1.49*** (-0.7) | **1.17*** (-0.55) | -0.09 (-0.4) | 0.18 (-0.95) | 0.79 (-0.79) |

**Supplementary Table 2b** Cognition/function ~ education + EPVS_WM_log + Group (AD vs. CO) + EPVS_WM_log*Group (AD vs. CO)

| Model - AD:CO | CDR-SoB *B* (Std. Error) | Memory *B* (Std. Error) | Executive *B* (Std. Error) | Visuospatial *B* (Std. Error) | Language *B* (Std. Error) |
|---|---|---|---|---|---|
| Intercept | -1.63 (-1.99) | 1.34 (-1.88) | **-2.71*** (-1.29) | 2.17 (-3.22) | 0.85 (-2.63) |
| Education | 0 (-0.06) | -0.02 (-0.05) | 0.02 (-0.04) | -0.14 (-0.09) | -0.08 (-0.08) |
| EPVS_WM_log | **-1.55**** (-0.51) | **0.89*** (-0.41) | -0.28 (-0.28) | 0.22 (-0.7) | 0.44 (-0.57) |
| Group: CO | 1.71 (-2.43) | -1.62 (-1.88) | 1.9 (-1.36) | 0.55 (-3.2) | -0.54 (-2.69) |
| EPVS_WM_log*Group: CO | **1.57*** (-0.72) | **-1.22*** (-0.56) | 0.07 (-0.4) | -0.2 (-0.95) | -0.87 (-0.8) |

**Supplementary Table 2c** Cognition/function ~ education + EPVS_WM_log + Group (EOAD vs. LOAD) + EPVS_WM_log*Group (EOAD vs. LOAD)

| Model - LOAD:EOAD | CDR-SoB *B* (Std. Error) | Memory *B* (Std. Error) | Executive *B* (Std. Error) | Visuospatial *B* (Std. Error) | Language *B* (Std. Error) |
|---|---|---|---|---|---|
| Intercept | 6.19 (-4.18) | -1.7 (-2.42) | -2.57 (-2.15) | 1.04 (-5.83) | -1.42 (-3.82) |
| Education | -0.03 (-0.08) | -0.01 (-0.06) | 0.1 (-0.05) | -0.03 (-0.15) | -0.03 (-0.1) |
| EPVS_WM_log | 0.38 (-1.16) | 0.22 (-0.64) | 0.36 (-0.57) | 1.17 (-1.55) | 0.11 (-1.02) |
| Group: LOAD | -7.03 (-5.01) | 2.54 (-3.03) | -1.91 (-2.47) | -1.47 (-7.24) | 0.74 (-4.69) |
| EPVS_WM_log*Group: LOAD | -1.87 (-1.44) | 0.61 (-0.87) | -0.73 (-0.71) | -1.18 (-2.07) | 0.18 (-1.34) |

**Supplementary Table 2d** Cognition/function ~ age + education + EPVS_WM_log + Group (CO vs. LOAD) + EPVS_WM_log*Group (CO vs. LOAD)

| Model - LOAD:CO | CDR-SoB *B* (Std. Error) | Memory *B* (Std. Error) | Executive *B* (Std. Error) | Visuospatial *B* (Std. Error) | Language *B* (Std. Error) |
|---|---|---|---|---|---|
| Intercept | -0.7 (-2.18) | -1.74 (-1.58) | -0.23 (-1.34) | 1.68 (-2.79) | 1.4 (-2.34) |
| Age | 0.01 (-0.02) | 0.02* (-0.01) | -0.01 (-0.01) | 0.02 (-0.02) | -0.02 (-0.02) |
| Education | 0 (-0.06) | -0.02 (-0.05) | 0.02 (-0.04) | -0.14 (-0.09) | -0.08 (-0.08) |
| EPVS_WM_log | 0.02 (-0.49) | -0.25 (-0.36) | -0.22 (-0.29) | 0.08 (-0.64) | -0.5 (-0.54) |
| Group: LOAD | -1.45 (-2.38) | 1.15 (-1.84) | -1.92 (-1.36) | -0.84 (-3.23) | 0.5 (-2.67) |
| EPVS_WM_log*Group: LOAD | **-1.51*** (-0.7) | **1.11*** (-0.54) | -0.07 (-0.4) | 0.12 (-0.96) | 0.87 (-0.79) |

***p<0.001; **p<0.01; *p<0.05

**Supplementary Table 2e** Cognition/function ~ age + education + EPVS_WM_log + Group (AD vs. CO) + EPVS_WM_log*Group (AD vs. CO)

| Model - AD:CO | CDR-SoB<br>*B* (Std. Error) | Memory<br>*B* (Std. Error) | Executive<br>*B* (Std. Error) | Visuospatial<br>*B* (Std. Error) | Language<br>*B* (Std. Error) |
|---|---|---|---|---|---|
| Intercept | -2.31<br>(-2.38) | -0.45<br>(-2.02) | -2.1<br>(-1.51) | 0.86<br>(-3.54) | 2.19<br>(-2.94) |
| Age | 0.01<br>(-0.02) | 0.02*<br>(-0.01) | -0.01<br>(-0.01) | 0.02<br>(-0.02) | -0.02<br>(-0.02) |
| Education | 0<br>(-0.06) | -0.02<br>(-0.05) | 0.02<br>(-0.04) | -0.14<br>(-0.09) | -0.09<br>(-0.08) |
| EPVS_WM_log | **-1.56****<br>(-0.51) | **0.91***<br>(-0.4) | -0.27<br>(-0.28) | 0.22<br>(-0.7) | 0.45<br>(-0.57) |
| Group: CO | 1.78<br>(-2.44) | -1.31<br>(-1.84) | 1.83<br>(-1.37) | 0.81<br>(-3.22) | -0.82<br>(-2.7) |
| EPVS_WM_log*Group: CO | **1.58***<br>(-0.72) | **-1.15***<br>(-0.55) | 0.05<br>(-0.41) | -0.13<br>(-0.95) | -0.94<br>(-0.8) |

***p<0.001; **p<0.01; *p<0.05

**Supplementary Table 2f** Cognition/function ~ age + education + EPVS_WM_log + Group (EOAD vs. LOAD) + EPVS_WM_log*Group (EOAD vs. LOAD)

| Model - LOAD:EOAD | CDR-SoB<br>*B* (Std. Error) | Memory<br>*B* (Std. Error) | Executive<br>*B* (Std. Error) | Visuospatial<br>*B* (Std. Error) | Language<br>*B* (Std. Error) |
|---|---|---|---|---|---|
| Intercept | 6.04<br>(-5.64) | -1.39<br>(-3.37) | -2.63<br>(-2.86) | -6.52<br>(-7.89) | -0.84<br>(-5.19) |
| Age | 0<br>(-0.06) | 0<br>(-0.03) | 0<br>(-0.03) | 0.11<br>(-0.08) | -0.01<br>(-0.05) |
| Education | -0.03<br>(-0.08) | -0.01<br>(-0.06) | 0.1<br>(-0.06) | -0.01<br>(-0.15) | -0.03<br>(-0.1) |
| EPVS_WM_log | 0.38<br>(-1.17) | 0.22<br>(-0.65) | 0.36<br>(-0.58) | 1.03<br>(-1.54) | 0.12<br>(-1.03) |
| Group: LOAD | -7.07<br>(-5.12) | 2.58<br>(-3.07) | -1.93<br>(-2.55) | -2.85<br>(-7.26) | 0.87<br>(-4.79) |
| EPVS_WM_log*Group: LOAD | -1.87<br>(-1.45) | 0.6<br>(-0.88) | -0.73<br>(-0.72) | -1.04<br>(-2.06) | 0.17<br>(-1.35) |

***p<0.001; **p<0.01; *p<0.05

**Supplementary Table 3**

PET marker ~ age + education + EPVS_WM_log + CDR-Sum of Boxes + Group(EOAD_LOAD) + EPVS_WM_log *CDR-Sum of Boxes *Group (EOAD_LOAD)

| | Temporal meta ROI FTP | Global cortical FTP | Precuneus FTP | PIB Centiloids |
|---|---|---|---|---|
| | *B* (Std. Error) | *B* (Std. Error) | *B* (Std. Error) | *B* (Std. Error) |
| Intercept | **5.35 **** (-1.84) | **4.35 **** (-1.4) | **6.41 **** (-2.36) | -7.12 (-166.45) |
| Age | -0.01 (-0.01) | -0.02 (-0.01) | -0.03 * (-0.01) | 0.81 (-1.05) |
| Education | 0 (-0.02) | | | |
| EPVS_WM_log | 0.87 (-0.5) | 0.56 (-0.38) | 0.75 (-0.63) | -6.29 (-44.53) |
| CDR-Sum of Boxes | **-0.71 *** (-0.35) | -0.5 (-0.26) | -0.67 (-0.44) | -11.96 (-30.96) |
| Group: LOAD | -3.25 (-2.12) | -2.06 (-1.6) | -2.69 (-2.69) | -129.34 (-190.88) |
| EPVS_WM_log * Boxscore | **-0.24 *** (-0.1) | **-0.17 *** (-0.08) | -0.23 (-0.13) | -4.86 (-9.09) |
| EPVS_WM_log * Group:LOAD | -0.94 (-0.61) | -0.58 (-0.46) | -0.74 (-0.78) | -27.29 (-54.94) |
| CDR-Sum of Boxes * Group:LOAD | **1.16 **** (-0.44) | **0.81 *** (-0.33) | 1.1 (-0.56) | 76.94 (-39.79) |
| EPVS_WM_log * CDR-SoB*Group:LOAD | **0.34 **** (-0.13) | **0.24 *** (-0.09) | 0.32 (-0.16) | 20.92 (-11.31) |

**Supplementary Figure 1.** Including the entire cohort (both Controls and AD), EPVS volume only weakly correlates with white matter volume.

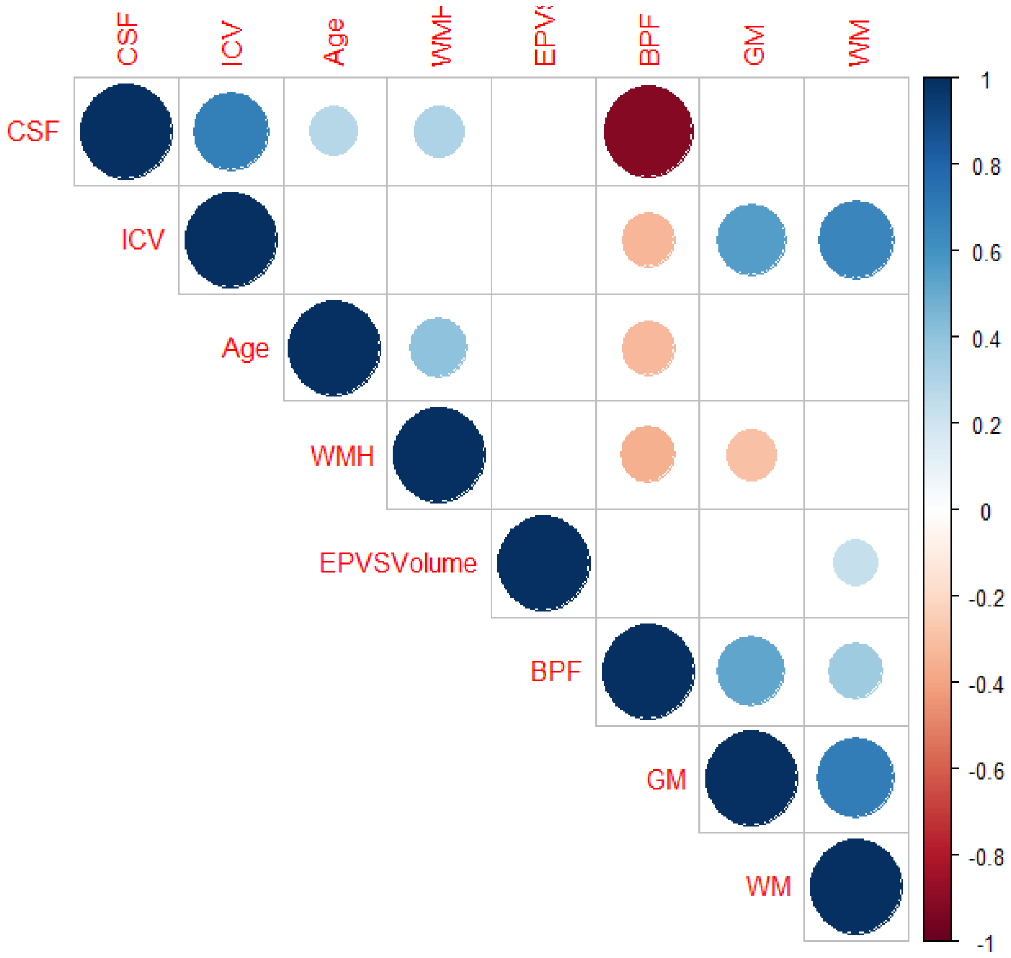

**Supplementary Figure 2.** EPVS volume correlation with age, gray matter volume, white matter volume, white matter hyperintensity log transformed, brain parenchymal fraction, by disease group.

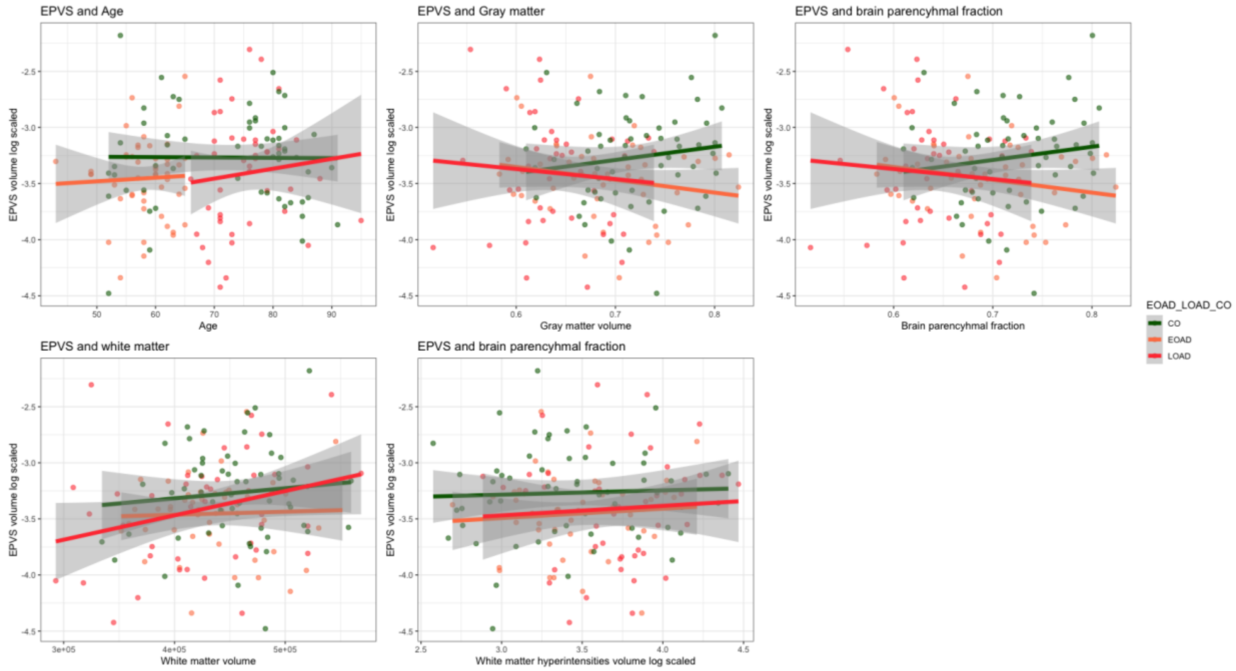

**Supplementary Figure 3.** EPVS relationship with function and cognition (memory, executive, visuospatial, language) in EOAD, LOAD and CO.

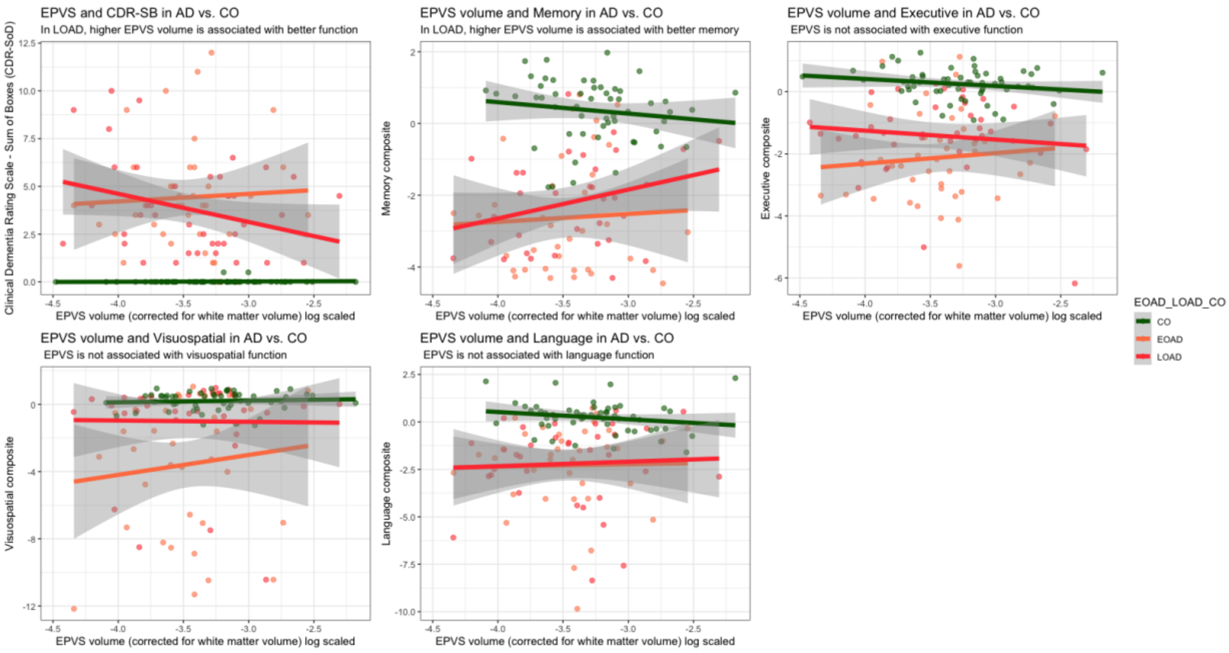

**Supplementary Figure 4.** Three-way interaction of tau by disease group (EOAD vs LOAD) by log transformed EPVS volume (lower tertile in red, middle tertile in blue, and highest tertile in green) on functional performance as measured by clinical dementia rating scale sum of boxes. In LOAD, patients in the highest tertile of EPVS volume (green) show lowest CDR scores for the same amount of tau as in the middle and lowest EPVS volume tertiles (blue and red). This model was not statistically significant.

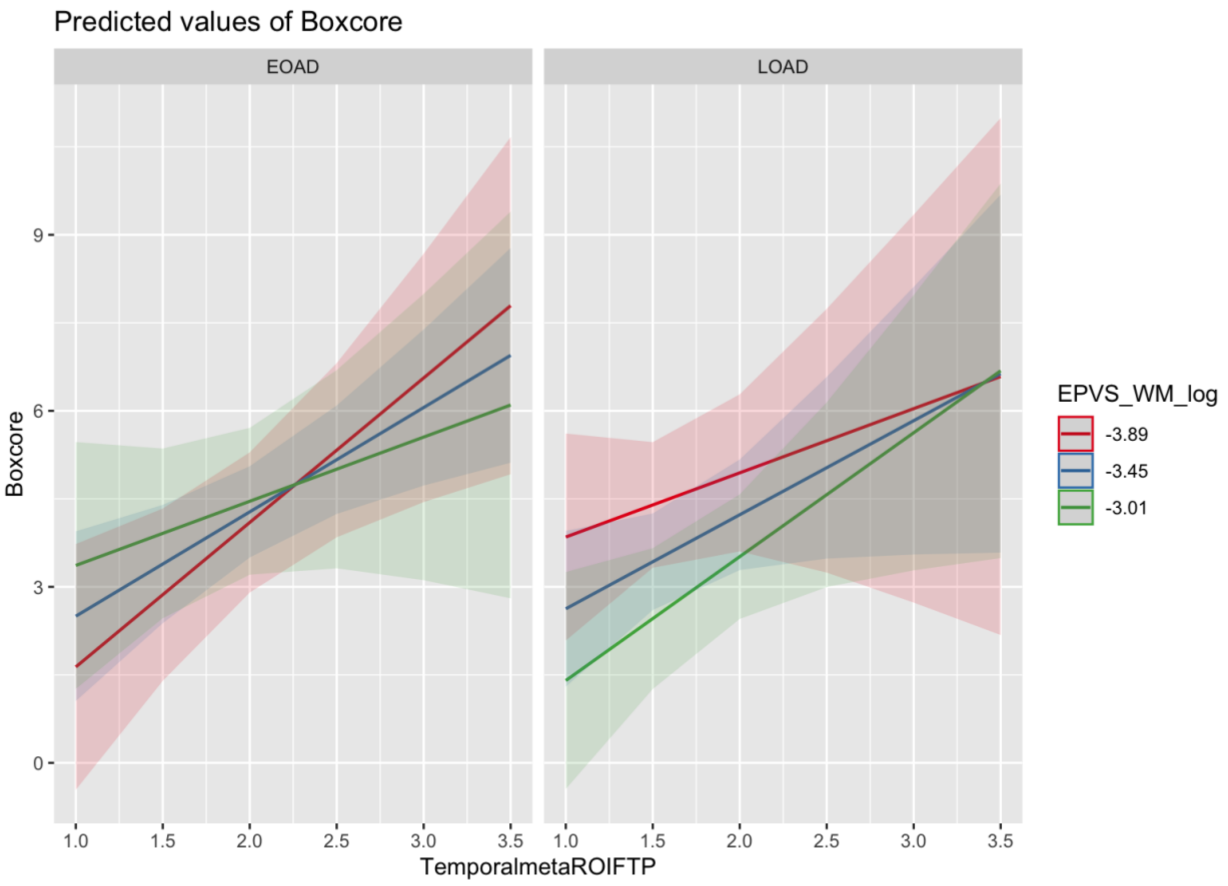
